# Incidence and Factors Associated with Visual Axis Opacification Following Pediatric Cataract Surgery at KCMC from 2013 to 2023

**DOI:** 10.1101/2025.08.01.25332692

**Authors:** Shibu Juma Nkuwi, Sara Kweka, Andrew Makupa, Furahini Mndeme

## Abstract

**Background:** Pediatric cataract is a significant cause of childhood blindness globally, with visual axis opacification (VAO) being a leading and challenging postoperative complication.

**Objective:** To evaluate the incidence and factors associated with visual axis opacification (VAO) following pediatric cataract surgery at KCMC Eye Department from January 2013 to January. 2023.

**Methods:** A retrospective cohort study of 345 children (612 eyes) who underwent cataract surgery was conducted. Demographic and clinical data were collected from patient records. Data analysis was performed using STATA version 17. Kaplan-Meier survival curves assessed the probability of VAO over time. A Poisson regression model was used to identify factors associated with VAO, with significance set at P < 0.05.

**Results:** Among 345 children, 189 (54.8%) were male, and 267 (77.4%) had bilateral cataracts. The median age at surgery was 28.5 months (range 9-72). Intraocular lenses were implanted in 519 (84.8%) eyes. The overall incidence of VAO was 19.9% (122/612), with an incidence rate of 0.251 per year. Secondary surgeries were performed in 20.3% (124/612) of eyes, mainly to clear the visual axis. Postoperatively, 69.4% of eyes had no visual impairment. Significant factors associated with VAO included age at surgery <60 months (AHR = 4.90; 95% CI: 2.77-8.70); P-value<0.001), surgical technique (LWO+IOL) (AHR = 7.58; 95% CI: 3.85-14.91); P-value<0.001), and postoperative acute fibrinous reaction (AHR = 5.91; 95% CI: 4.01-8.71); P-value<0.001).

**Conclusion:** The incidence of VAO at KCMC is consistent with global data. Early age at surgery, surgical technique without PPC and AV, and postoperative inflammation were significantly associated with VAO. Adoption of preventive strategies and enhanced postoperative care are critical for improving visual outcomes.

## INTRODUCTION

Pediatric cataract remains one of the leading treatable causes of childhood blindness globally (1). Its prevalence is estimated between 0.32 and 22.9 per 10,000 children worldwide. In Africa, over 19,000 new cases are reported annually (2). In Tanzania, the prevalence of severe visual impairment and blindness stands at 0.05%, with lens-related conditions accounting for 27% of the causes (3).

Pediatric cataract is defined as opacification of the natural crystalline lens, either present at birth or developing later in childhood. It can be unilateral or bilateral, with etiologies that include idiopathic, hereditary, infectious, metabolic, or part of syndrome (4). Since visual system continues to mature after birth, any lens opacity may interrupt this critical developmental process, possibly resulting in irreversible visual impairment or blindness (5). Such impairment imposes emotional, social, and economic burdens on children, families, and society, prompting global initiatives such as VISION 2020 to prioritize congenital cataracts (6).

Surgical intervention is the mainstay for visually significant pediatric cataracts. While surgical techniques have evolved, especially in low resourced regions, the standard approach of primary posterior capsulotomy with in -the-bag intraocular lens (IOL) implantation and anterior vitrectomy remains widely considered (7). Despite advancements, Visual Axis Opacification (VAO) continues to be a major postoperative complication, with incidence rates reported between 7.5% and over 50% globally, even in developed healthcare systems (8)(9). VAO is largely due to rapid proliferation of lens epithelial cells and fibrous tissue growth on the anterior vitreous face and residual capsule (10).

VAO can compromise initially successful surgical outcomes. And presence of VAO often necessitates secondary surgeries under general anesthesia, this raise concerns about additional risk, especially neurodevelopment effects in young children (11). The resulting visual axis obstruction can deprive the visual system of required stimuli, contributing to amblyopia, which underscores the need for timely detection and management (12).

Secondary surgical interventions are commonly reported following pediatric cataract surgery. These include capsulotomy and membranectomy to restore a clear visual axis, as well as IOL repositioning or secondary IOL implantation (13)(14). Visual outcomes after these procedures have shown significant improvement (15). Several risk factors for VAO have been identified, including young age at surgery, microphthalmia, persistent hyperplastic fetal vasculature (PHFV), use of intraocular lenses, IOL design and material, position of fixation, and the intensity of postoperative steroid therapy (16).

At Kilimanjaro Christian Medical Centre (KCMC), a tertiary referral hospital in northern Tanzania, pediatric cataract surgeries have been routinely performed for over a decade by experienced ophthalmologists. Despite this experience, local data on the incidence and contributing factors of VAO remains limited.

This study aims to determine the incidence and associated factors of VAO following pediatric cataract surgery at KCMC over a 10 year period. The findings will contribute to understanding the scope of the problem in our setting, help assess current practices, and support the development of targeted strategies to prevent or manage VAO, particularly in resource limited countries including Tanzania.

## METHODOLOGY

### Study Design and Setting

This was a retrospective cohort study conducted at the Ophthalmology Department of Kilimanjaro Christian Medical Centre (KCMC), a zonal referral hospital located in Moshi Municipality, Kilimanjaro Region, Tanzania. The study included 345 children who underwent congenital or developmental cataract surgery at KCMC between January 2013 and January 2023. All patients were followed up for one year post operatively.

### Ethical Approval

Ethical clearance was obtained from the Kilimanjaro Christian Medical University College **(KCMUCo) Research** and **Ethics Committee** with the following **number (Ref: PG. 30/2023)**. This study was **approved on 15**^**th**^ **August 2023** with duration of one year from **15**^**th**^ **August 2023 to 14**^**th**^ **August 2024**. All data were collected with the study period given and were **anonymized** and treated with **strict confidentiality**.

### Participants

The study included children under 16 years of age who were diagnosed with congenital or developmental cataracts and underwent surgery at KCMC within the study period. Exclusion criteria were children diagnosed with traumatic or uveitic cataracts, incomplete or missing demographic and clinical data also cases undergoing only secondary intraocular lens (IOL) implantation during the study period. A consecutive non probability sampling technique was used.

### Data Collection

Medical records were reviewed retrospectively using a structured data collection tool. The following data were extracted, demographic information, clinical characteristics such as pre- and post-operative best corrected visual acuity, intraocular pressure, surgical details including type and power of IOL, surgical technique, postoperative outcomes and complications, secondary surgeries and their indications.

### Statistical Analysis

Data analysis was performed using STATA version 17 (StataCorp LLC, College Station, Texas, USA). Categorical variables were summarized using frequencies and percentages. Numerical variables were presented as means with standard deviation (SD) or medians with interquartile range (IQR), as appropriate. Kaplan-Meier survival curves were used to estimate the survival probability of eyes without visual axis opacification (VAO) over time. Stratified analyses were performed by age at surgery and surgical technique used. Group comparisons were conducted using the log-rank test, with significance set at *P* < 0.05.

A Poisson regression model was employed to identify factors associated with VAO. Univariate analyses were conducted to estimate crude hazard ratios (CHR), followed by multivariable analysis to obtain adjusted hazard ratios (AHR) with corresponding 95% confidence intervals (CI). Variables with *P* < 0.05 were considered statistically significant.

## RESULTS

This study included a total of 345 (612 eyes) participants with median age at surgery of 28.5*(IQR: 9-72)* months. Among them, 189 (54.8%) were male and 267 (77.4%) had bilateral cataract while the majority of participants 231 (67%), originated from the Northern zone. Out of 612 operated eyes, 304 (49.7%) underwent surgery at ≤24 months of age. The most common presenting complaints were a combination of white pupillary reflex, strabismus, and poor vision in 244 (39.8%) eyes. The median time from symptom onset to hospital presentation was 12 months *(IQR: 5–36)*, and the median wait time from hospital diagnosis to surgery was 5 days (IQR: 2–8). Commonly used surgical technique was LWO+PPC+AV+BAG IOL in 343 (56%) eyes. At one year follow up 519 eyes (84.8%) were pseudophakic and 93 (15.2%) aphakic. A commonly prescribed post operative topical drug was a combination of dexamethasone and chloramphenicol in 534 (87.3%) eyes. (**Table 1)**

**Table 1.** Demographic and Clinical characteristics of participants’ eyes underwent cataract surgery (N=612)

| **Variable** | **Frequency** | **Percentage** |
| --- | --- | --- |
| **Age at Surgery in months** |  |  |
| ≤24 | 304 | 49.7 |
| 25-60 | 127 | 20.8 |
| ≥61 | 181 | 29.6 |
| *Median (IQR)* | *28.5(9-72)* |  |
| **Sex of the participant** |  |  |
| Male | 189 | 54.8 |
| Female | 156 | 45.2 |
| **Laterality of cataract** |  |  |
| Unilateral | 78 | 22.6 |
| Bilateral | 267 | 77.4 |
| **Waiting time for surgery in days** |  |  |
| *Median (IQR)* | *5(2-8)* |  |
| **Time from symptom onset to hospital presentation (months)** |  |  |
| *Median (IQR)* | *12(5-36)* |  |
| **IOL power implanted in Diopters** |  |  |
| *Median (IQR)* | *25(22-28)* |  |
| **Presenting symptoms/complain** |  |  |
| White pupillary reflex | 153 | 25 |
| Poor Vision | 214 | 35 |
| Strabismus | 1 | 0.2 |
| Combined symptoms | 244 | 39.8 |
| **Surgical Approach Used** |  |  |
| Anterior approach | 561 | 91.7 |
| Pars Plana approach | 51 | 8.3 |
| **Surgical Technique used** |  |  |
| LWO+PPC+AV+BAG IOL | 343 | 56 |
| LWO+PPC+AV+SULCUS IOL | 98 | 16 |
| LWO+BAG IOL | 78 | 12.8 |
| LWO+PPC+AV+APHAKIA | 93 | 15.2 |
| **Post Op. Lens status at 1 year of FU** |  |  |
| Pseudophakia | 519 | 84.8 |
| Aphakia | 93 | 15.2 |
| **Post Op. Topical steroid used** |  |  |
| Dexamethasone + Chloramphenicol | 534 | 87.3 |
| Prednisolone acetate 1% | 78 | 12.7 |

In this study the overall incidence of VAO was 19.9% (122/612), with a calculated incidence rate of 25.1% per year. The median follow-up period was 1 year (IQR: 0.36–1.11).Kaplan-Meier survival analysis revealed overall survival probability of (0.8) while age at surgery ≤24 months and LWO+IOL surgical technique had lower survival probability of 0.75 and 0.65 when compared to other age groups and surgical techniques respectively. **((Figure 1 (A), (B) and (C))**

**Figure 1.**
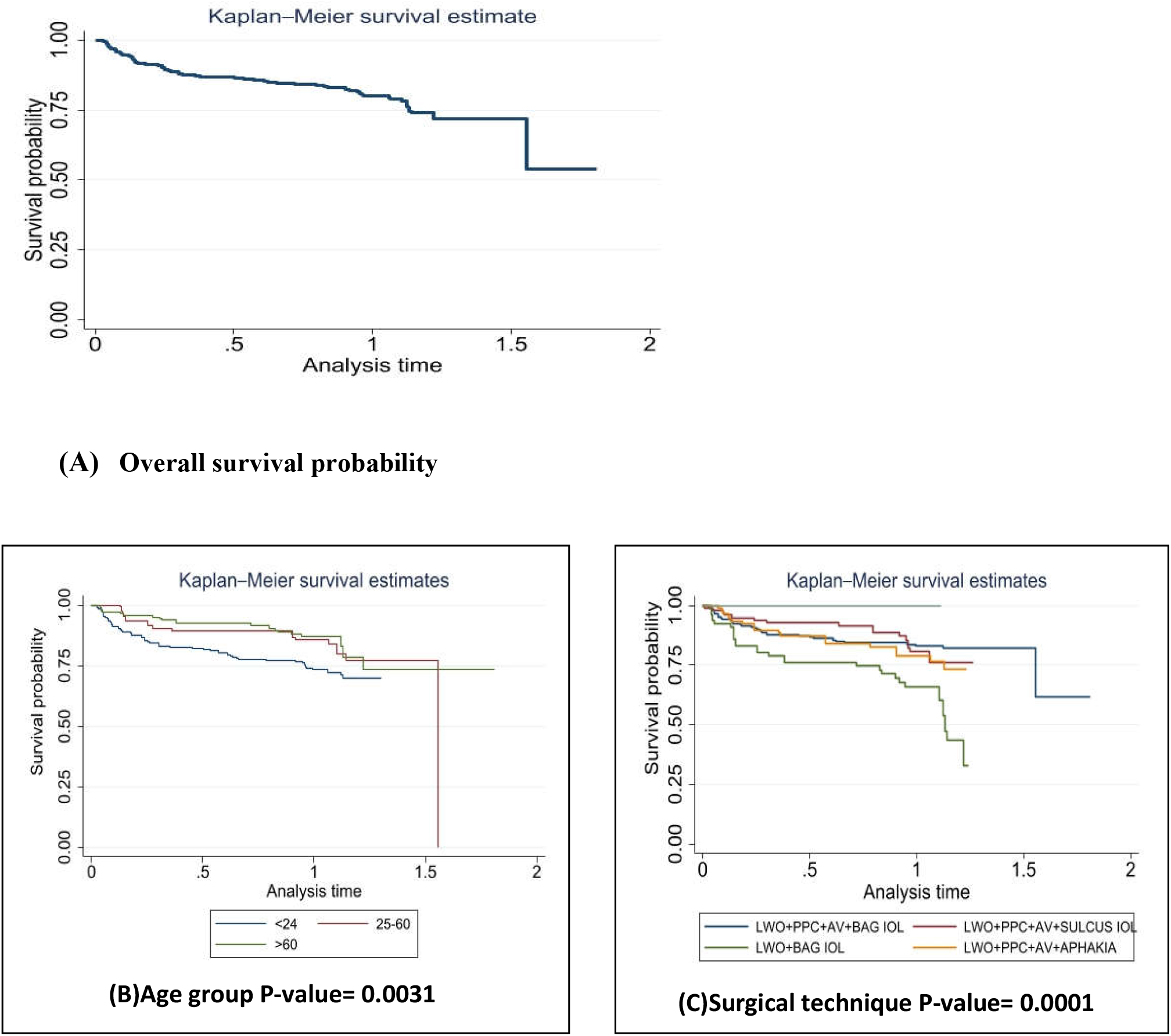
(A, B&C) demonstrate Kaplan Meier survival curves showing overall survival probability and the survival probability by age groups and surgical techniques used.

In our study a number of eyes needed secondary surgeries in which a total of 124 eyes (20.3%) required secondary surgical interventions, yielding an incidence rate 1.72 per year. The most frequent procedures included surgical capsulotomy (10.3%), membranectomy (6.4%), and YAG laser capsulotomy (3.6%). Other interventions done included secondary IOL insertion (3.1%), IOL reposition (1.3%), IOL exchange (0.7%), synechiolysis (1.8%), and pupilloplasty (2.0%) **(Table 2)**

**Table 2.** Secondary surgeries performed following pediatric cataract surgery (N=612)

| **Variable** | **Frequency** | **Percentage** |
| --- | --- | --- |
| Surgical Capsulotomy | 63 | 10.3 |
| Membranectomy | 39 | 6.4 |
| IOL reposition | 8 | 1.3 |
| Secondary IOL insertion | 19 | 3.1 |
| YAG capsulotomy | 22 | 3.6 |
| IOL exchange | 4 | 0.7 |
| Synechiolysis | 11 | 1.8 |
| Pupilloplasty | 12 | 2 |

Concerning visual outcome in this study, one year post-operatively, 69.4% of eyes had no visual impairment, 17% had moderate, and 4.6% severe and 9% remained blind. This demonstrates a significant improvement in visual function following surgery compared to preoperative, 53.3% of eyes were classified as blind according to WHO criteria, 42.1% had moderate visual impairment, 3.5% severe visual impairment, and 1.2% had no impairment (**Figure 2**)

**Figure 2.**
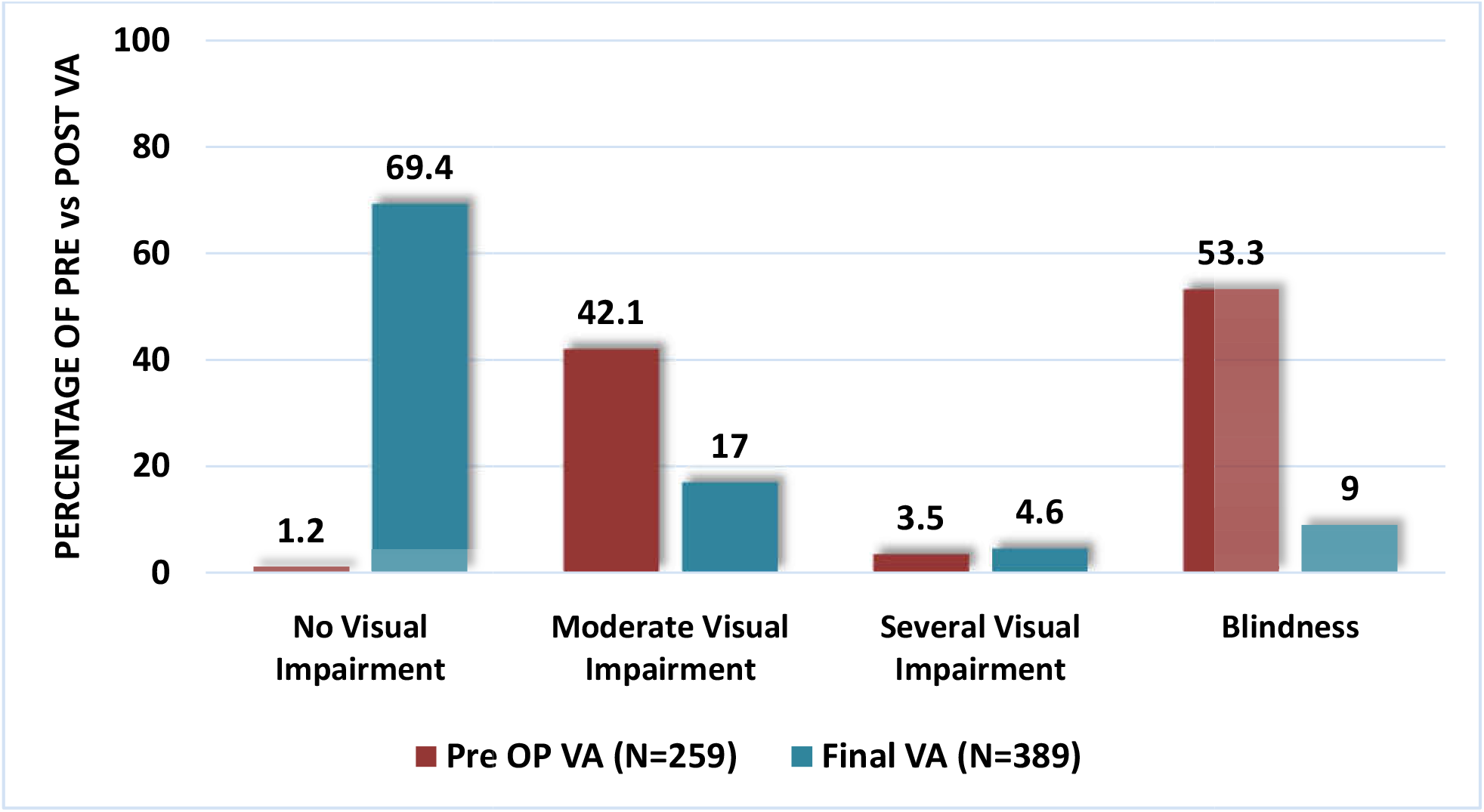
Proportion of Pre and Post operative best corrected visual acuity according to W.H.O classification.

In assessing factors associated with visual axis opacification univariate analysis showed that age at surgery <60 months (Age <60 months: CHR = 1.77 (95% CI: 1.14–2.76), P = 0.011), LWO+BAG IOL surgical technique (LWO+BAG IOL technique: CHR = 2.18 (95% CI: 1.22– 3.89), P < 0.001), and postoperative acute fibrinous reaction (Postoperative fibrinous reaction: CHR = 5.44 (95% CI: 3.75–7.90), P < 0.001) were significantly associated with VAO. In multivariable analysis: Age <60 months: AHR = 4.90 (95% CI: 2.77–8.70), P < 0.001), LWO+BAG IOL technique: AHR = 7.58 (95% CI: 3.85–14.91), P < 0.001) and postoperative fibrinous reaction: AHR = 5.91 (95% CI: 4.01–8.71), P < 0.001 were still significantly associated with VAO. Other variables such as IOL type, implantation site, hemoglobin levels, and steroid type did not show statistically significant associations with VAO. (**Table 3**)

**Table 3.** Factors Associated with Visual Axis Opacification occurrence following pediatric cataract surgery.

| Variable | CHR(95% CI) | P-value | AHR(95%CI) | P-value |
| --- | --- | --- | --- | --- |
| **Age at Surgery in months** |  |  |  |  |
| 60+ | Ref |  |  |  |
| <60 | 1.77(1.14-2.76) | 0.011 | 4.90(2.77-8.70) | <0.001 |
| **Post op. Lens status** |  |  |  |  |
| Aphakia | Ref |  |  |  |
| Pseudophakia | 0.98(0.60-1.62) | 0.945 |  |  |
| **Surgical technique used** |  |  |  |  |
| LWO+PPC+AV+APHAKIA | Ref |  | Ref |  |
| LWO+PPC+AV+BAG IOL | 0.78(0.46-1.32) | 0.359 | 0.81(0.47-1.38) | 0.441 |
| LWO+PPC+AV+SULCUS IOL | 0.82(0.42-1.60) | 0.843 | 0.78(0.40-1.53) | 0.472 |
| LWO+BAG IOL | 2.18(1.22-3.89) | <0.001 | 7.58(3.85-14.91) | <0.001 |
| **IOL model used** |  |  |  |  |
| Aphakia | Ref |  |  |  |
| ALCON SA60AT | 0.75(0.41-1.38) | 0.355 |  |  |
| ALCON MA60AC | 0.65(0.24-1.22) | 0.185 |  |  |
| SPNT200 | 1.68(0.98-2.90) | 0.062 |  |  |
| **IOL fixation site** |  |  |  |  |
| Aphakia | Ref |  |  |  |
| Bag IOL fixation | 1.09(0.64-1.86) | 0.752 |  |  |
| Sulcus IOL fixation | 0.88(0.45-1.73) | 0.721 |  |  |
| **IOL implantation** |  |  |  |  |
| Aphakia | Ref |  |  |  |
| Primary IOL implantation | 1.06(0.59-1.88) | 0.854 |  |  |
| Secondary IOL implantation | 1.45(0.52-4.07) | 0.479 |  |  |
| **Post op. Acute Fibrinous Reaction** |  |  |  |  |
| No | Ref |  | Ref |  |
| Yes | 5.44(3.75-7.90) | <0.001 | 5.91(4.01-8.71) | <0.001 |
| **Post Op. Topical steroid used** |  |  |  |  |
| Dexamethasone+chloramphenicol | Ref |  |  |  |
| Prednisolone acetate 1% | 1.43(0.89-2.32) | 0.140 |  |  |
| **Hemoglobin Level at time of surgery (g/dl)** |  |  |  |  |
| Anemia | Ref |  |  |  |
| Normal Hb level | 0.72(0.50-1.02) | 0.065 |  |  |

## DISCUSSION

This study included 345 children with congenital or developmental cataracts who underwent surgery at KCMC between 2013 and 2023. Most were male (54.8%), with a median surgery age of 28.5 months. Bilateral cataracts were more common (77.4%), and 612 eyes were analyzed with a median follow up time of one year.

In this study the overall incidence of visual axis opacification (VAO) was 19.9%, with an annual incidence rate of 25.1%. Our findings reflect the benefit of standard surgical protocols, including primary posterior capsulotomy (PPC) and anterior vitrectomy (AV). This aligns with rates reported in Kenya 17.8% (17) and 21.05% (18). However, the Kenyan study had significant loss to follow up and a shorter duration of follow up which might potentially underestimating the true incidence. In contrast, higher VAO incidences were reported in studies done in Sweden and United Kingdom 44.1% and 45% respectively (19) (20). The younger age group and longer follow up period in these studies likely contributed to the higher rates observed. A lower rate in Japan of 7.1% may reflect consistent PPC and AV use, though small sample size may have influenced the result (21).

In this study secondary surgery was required in 20.3% of eyes, mainly for VAO management, with surgical capsulotomy (10.3%) being the most common which aligning with findings from Tanzania (22), who reported a rate of 4.6%. In India, (23) found membranectomy to be the most common secondary intervention (6.9%). These observations highlight VAO as a leading cause of secondary procedures. A similar secondary surgery rate of 17.6% was reported in a Brazilian study (14), while a study from Bangladesh reported a higher rate of 33% (24), possibly due to a longer follow-up period of five years. Additional procedures in our study included synechiolysis (1.8%) and pupilloplasty (2.0%) further illustrating the need for refined surgical planning and long term management.

This study showed marked improvement in best corrected visual acuity post operatively in which at one year, 69.4% had no visual impairment while preoperatively 53.3% of eyes were blind. Improvements can be attributed to timely intervention, use of microsurgical techniques, intensive postoperative care, refractive errors correction, and amblyopia management by experienced surgeons in this study. Comparable outcomes were reported in England where 73.5% had visual acuity of 6/12 or better (25). Visual outcomes in this study were better than those reported in Kenya where no visual impairment accounted to (44%, 44.5%) (18), (26) and Bangladesh (48%) (24), where shorter follow-up durations may have limited observed visual improvement in these studies.

Our study identified risk factors for VAO to be age at surgery <60 months (AHR = 4.90, P < 0.001), surgical technique without PPC and AV (AHR = 7.58, P < 0.001), and postoperative fibrinous reaction (AHR = 5.91, P < 0.001). The association could be explained by vulnerability of younger age patient to high lens epithelial cells proliferation, strong inflammatory responses and prolonged healing process in younger age. Similarly increased risk of VAO in a younger age was also found in studies done by (27) (14) (13) in which VAO was noted especially in age less than 2 years. The association of surgical technique without PPC and AV with VAO could be explained by protective effect of doing primary posterior surgical capsulotomy and anterior vitrectomy in pediatric cataract as they reduce site for proliferation, migration and lodging of lens epithelial cells which are responsible for VAO. Findings in this study also corroborates with findings in study done by (21)(28) which also highlighted the efficiency of primary posterior capsulotomy and anterior vitrectomy in reducing VAO incidence following pediatric cataract surgery. Despite microsurgical technique use and intensive post-operative topical steroid use in this study post-operative acute fibrinous reaction proportion was found to be 13.2% and strongly associated with VAO occurrence this could be due to heightened inflammatory response in pediatric age group. Similar findings also have been reported in different studies (15)(22) These findings highlight the importance of intensive post-operative inflammatory control following pediatric cataract surgery. All IOLs were hydrophobic acrylic and univariate analysis suggested SPNT200 may have a higher VAO risk, while prior studies show hydrophilic IOLs and single-piece designs increase VAO risk (8) the differences in design and follow-up may explain discrepancies.

## CONCLUSION

This study provides critical insights into pediatric cataract surgery outcomes at a tertiary center in Northern Tanzania, representative of Sub-Saharan African settings. We observed a 19.9% incidence of visual axis opacification (VAO), with secondary surgery required in 20.3% of cases. Notably, surgical techniques incorporating posterior capsulotomy and anterior vitrectomy were associated with a reduced risk of VAO. Younger age at surgery, absence of PPC and AV, and postoperative acute fibrinous inflammation were significant risk factors. These findings highlight the importance of technique selection, vigilant postoperative care, and timely intervention to prevent VAO.

## Data Availability

all relevant data will be available under reasonable request from corresponding author

## RECOMMENDATIONS

We recommend consistent application of surgical protocols incorporating posterior capsulotomy and anterior vitrectomy, especially for children younger than 60 months. Enhanced training for pediatric ophthalmic surgeons in low-resource settings, robust postoperative inflammatory control, and caregiver education for follow-up compliance are crucial. Future prospective and multicenter studies are encouraged to explore long-term outcomes and modifiable risk factors for visual rehabilitation.

## ACKNOWLEDGEMENT

I thank Dr. Furahini Mndeme, Sara Kweka and Dr. Andrew Makupa for their invaluable mentorship and guidance.

I sincerely appreciate Kalvin Rwegoshora for his support in data analysis and biostatistics.

Finally, I am deeply grateful to my family for their unwavering encouragement and support throughout this journey.

